# Supporting virtual reality and active video game use in pediatric rehabilitation: Protocol for a mixed-methods feasibility study

**DOI:** 10.1101/2025.04.15.25325879

**Authors:** Audrey Ferron, Martin Lemay, Danielle E. Levac

## Abstract

**Background:** Virtual reality (VR) and active video game (AVG) systems that offer repetitive practice and multimodal feedback in engaging environments are attractive pediatric rehabilitation intervention options. Evidence supports their effectiveness to improve functional outcomes in multiple pediatric populations. However, VR/AVG integration into clinical practice faces multiple barriers, including limited access to these often-expensive technologies, a rapid development sector resulting in frequent obsolescence, and insufficient educational resources to help clinicians select appropriate games that match children’s therapeutic objectives. Knowledge translation initiatives that primarily target clinician knowledge and attitudes about VR/AVG use have shown limited success in facilitating adoption. To address these challenges, we used the Consolidated Framework of Implementation Research (CFIR) to structure development of the Technotheque, a multi-faceted VR/AVG support initiative at our large pediatric rehabilitation centre. The Technotheque addresses both access- and knowledge-based barriers to VR/AVG use via a dedicated gameplay space staffed by knowledge brokers who directly support clinicians in VR/AVG implementation in therapy sessions and provide education towards independent VR/AVG use.

**Objective:** To evaluate the feasibility of the Technotheque as a knowledge translation initiative to enhance VR/AVG use at our pediatric rehabilitation centre, as measured by demand, acceptability, adaptation, and implementation criteria.

**Methods:** Convergent mixed-methods design. We will use convenience and snowball sampling strategies to recruit clinicians (physiotherapists, occupational therapists, speech therapists, special educators, and neuropsychologists) to participate in this 4-month study. Following informed consent, participants will complete a modified ‘Assessing the Determinants of Prospective Take-up of Virtual Reality’ (ADOPT-VR2) instrument. Participants can then request individualized training and/or clinical implementation support with our knowledge brokers at a self-determined frequency and duration, and with their choice of clientele. Participants will be free to begin and end their study participation at any point during the four months. Data collection will include study-specific Technotheque pre-session objective and post-session feedback forms, a post-study ADOPT-VR reassessment, a CFIR-based satisfaction questionnaire, and individual semi-structured interviews. Quantitative analyses will examine demand (participant demographics, usage patterns, correlations between usage and ADOPT-VR2 scores), acceptability (satisfaction scores in relation to usage patterns and ADOPT-VR2 changes), adaptation (variety of professions, client populations, and clinical objectives), and implementation (pre-post ADOPT-VR2 changes and patterns of support needs over time). Qualitative data will be deductively analysed using the four feasibility criteria as a coding framework. Quantitative and qualitative results will be integrated to identify areas of alignment and/or divergence.

**Results:** Research Ethics Board approval has been obtained.

**Conclusions:** The Technotheque initiative is designed to address both access and knowledge barriers to VR/AVG use in pediatric rehabilitation. Study results will inform subsequent research efforts to evaluate the effectiveness of VR/AVG as a rehabilitation intervention and to examine the impact of this initiative on sustained VR/AVG use.

## Introduction

Pediatric rehabilitation is a multidisciplinary field in which professionals collaborate to help children with acquired or congenital disabilities address physical, cognitive, and psychosocial challenges and reach their full potential (1). In recent years, many rehabilitation disciplines have incorporated interactive technologies such as virtual reality (VR) within their intervention toolbox. VR is defined as “an artificial environment which is experienced through sensory stimuli (such as sights and sounds) provided by a computer, and in which one’s actions partially determine what happens in the environment” (2). This broad definition encompasses visually immersive technologies (three-dimensional virtual environments viewed through a head-mounted display) and non- or semi-immersive environments (two-dimensional virtual environments viewed on a flat-screen display). Semi-immersive visual displays, in which body movements tracked by cameras or sensors control interaction with the virtual environment, are often referred to as active video games (AVGs). VR/AVG systems may be custom developed for rehabilitation use (customized systems) or developed for the public (non-customized systems).

VR/AVGs are used to target physical, cognitive, and psychosocial goals in a variety of pediatric populations, although substantial research efforts explore their use with children with neuromotor impairments such as cerebral palsy (CP) and traumatic brain injury (3–6). Theoretical rationale for VR/AVG use lies in motor learning theory, with arguments that these interventions can target principles of repetition, ‘just-right’ challenge, augmented feedback, and motivation that improve skill learning and retention (7–10). Studies demonstrate that VR/AVG interventions can improve gross motor function, upper extremity use, balance and postural control (11–15). Exergaming, defined as the use of AVGs to promote physical activity, has been evaluated in children with autism spectrum disorder, CP and spina bifida (16,17), implemented in home-based interventions for youth with mobility challenges (18), and evaluated as a modality to enhance early mobility in the intensive care unit.(19) In cognitive and psychosocial rehabilitation, VR/AVG use can offer equitable social participation opportunities (20) and enable assessment of cognitive functions such as information processing speed and attention in ecological contexts, like simulated classroom environments (21,22).

Despite a growing evidence base, clinicians interested in using VR/AVGs in pediatric rehabilitation face substantial environmental and educational barriers. Knowledge gaps identified by clinicians include limited technical skills, lack of familiarity with available technologies, lack of allocated training time, and uncertainty about matching specific VR/AVG system affordances to individual client goals (23–28). (25,29). In addition, a lack of environmental and organizational support are critical barriers to VR/AVG adoption (25,26). Knowledge transfer initiatives fail to increase usage if environmental barriers are not addressed (30). For example, structural factors such as dedicated space, accessibility, inadequate Wi-Fi and restrictive firewalls, and insufficient technical support and funding can hinder implementation efforts (24,26,28,31,32).

Surveys in multiple countries demonstrate that there are varying degrees of VR/AVG adoption across rehabilitation contexts. In Canada, a 2016 survey found that approximately 46% of physiotherapists and occupational therapist respondents reported experience with VR/AVGs, but only 12% were current users (24). In the US, about 31% of clinician respondents reported current VR/AVG use in a 2017 survey(33). In Korea, 61% of respondents had VR/AVG experience (28), while in the UK only 7% of pediatric physiotherapists reported current VR use (27). In a 2024 European survey, 45.5% of respondents reported using VR/AVGs, with lack of training amongst the most frequently reported barriers to use (34). Among occupational therapists in Canada, 12.4% recommended technology use within rehabilitation for their elderly clients (35). Across these studies, non-customized systems (such as the Nintendo Wii/WiiFit and Microsoft’s Kinect for Xbox) were more commonly used than customized systems.

Initiatives to support the adoption and use of VR/AVGs in clinical practice include educational resources that address clinicians’ knowledge gaps, such as frameworks that classify VR/AVG games and provide decision-making suggestions (36–39). Studies have demonstrated that strategies focused on providing education in the form of clinical manuals or online resources (31,40) can change attitudes and boost self-reported confidence, but that hands-on practice sessions and technical support are essential ((31,41) (42). Integrated knowledge translation strategies that involve clinicians from the beginning, and ideally include a needs assessment process to identify and target site-specific facilitators and barriers, are recommended to ensure that selected strategies align with clinical realities (25,29). Dedicated personnel to support colleagues, such as a knowledge broker or a clinician champion, can facilitate VR/AVG adoption (26,41,43).

Given that accessibility is a recognized barrier, the creation of a dedicated VR/AVG play space within a rehabilitation centre can be advantageous (43). Nguyen et al. (2019) assessed the impact of a ‘VR exergaming’ room on VR use in a stroke rehabilitation center. In this context, they found that facilitators to VR use included the expert clinician who created individualized programs for clients with different functional abilities, client motivation to use VR, and the opportunity for VR room use to provide additional therapy hours outside of regular sessions. Barriers included staffing limitations, scheduling complexities and inadequate training in using VR in the absence of the clinician champion. The combination of institutional support, dedicated staffing, and comprehensive clinician training appears to be key for effective VR/AVG implementation in rehabilitation.

Guided by this knowledge base, we developed a multi-faceted VR/AVG implementation support initiative called the Technotheque at the Centre de readaptation Marie Enfant (CRME) in Montreal, QC. The Technotheque is a room dedicated to VR/AVG use and staffed with knowledge brokers: expert consultants who provide educational and clinical implementation assistance to CRME clinicians, with and without the presence of a pediatric client. The room has six gameplay stations hosting 12+ VR/AVG systems and 150+ games, all connected by a user-friendly centralized control system (**Figure 1**). We used the Consolidated Framework for Implementation Research (CFIR) to guide the design, implementation and evaluation of the Technotheque (44,45). The CFIR has 5 domains (intervention characteristics, outer setting, inner setting, characteristics of individuals, and process) that provide a comprehensive structure to assess the multiple factors that may influence implementation success. **Table 1** outlines how the 5 CFIR domains informed the Technotheque development process.

**Table 1:** Technotheque development activities targeted to implementation success, organized by CFIR domain.

| CFIR domain | Technotheque development activities |
| --- | --- |
| **Intervention Characteristics (VR/AVGs)** | • Playtesting and analysis of each VR/AVG game<br>• Development of a classification framework to categorize games based on different therapeutic requirements<br>• Creation of information sheets describing each game and its requirements to inform decision-making<br>• Creation of short video presentation examples of use |
| **Outer Setting (Marie Enfant Rehabilitation Centre [CRME])** | • Needs assessment to understand facilitators and barriers to current VR/AVG use at CRME |
| **Inner Setting (The Technotheque)** | • Identification of a dedicated room<br>• Availability of equipment (e.g., balance boards, weights, mats, adaptive controllers etc.) for adaptations and modifications<br>• Creation of a centralized screen control system to provide a single access point to each VR/AVG system<br>• Preliminary testing of Technotheque use in a restricted context (45)<br>• 3-month pilot phase with 9 clinicians and 13 children and youth |
| **Individual characteristics (Clinicians at CRME)** | • Creation of a committee of ‘super-users’ during the pilot phase to validate fit of Technotheque human and material resources with clinical needs |
| **Process (Design of study procedures)** | • Super-user committee piloted study procedures and feedback informed changes for the feasibility study<br>• Technotheque knowledge broker training<br>• Clinician study participation time (outside of clinical use with clients) paid for by research funds<br>• Administrative support provided for Technotheque reservations and study form completion |

**Figure 1.**
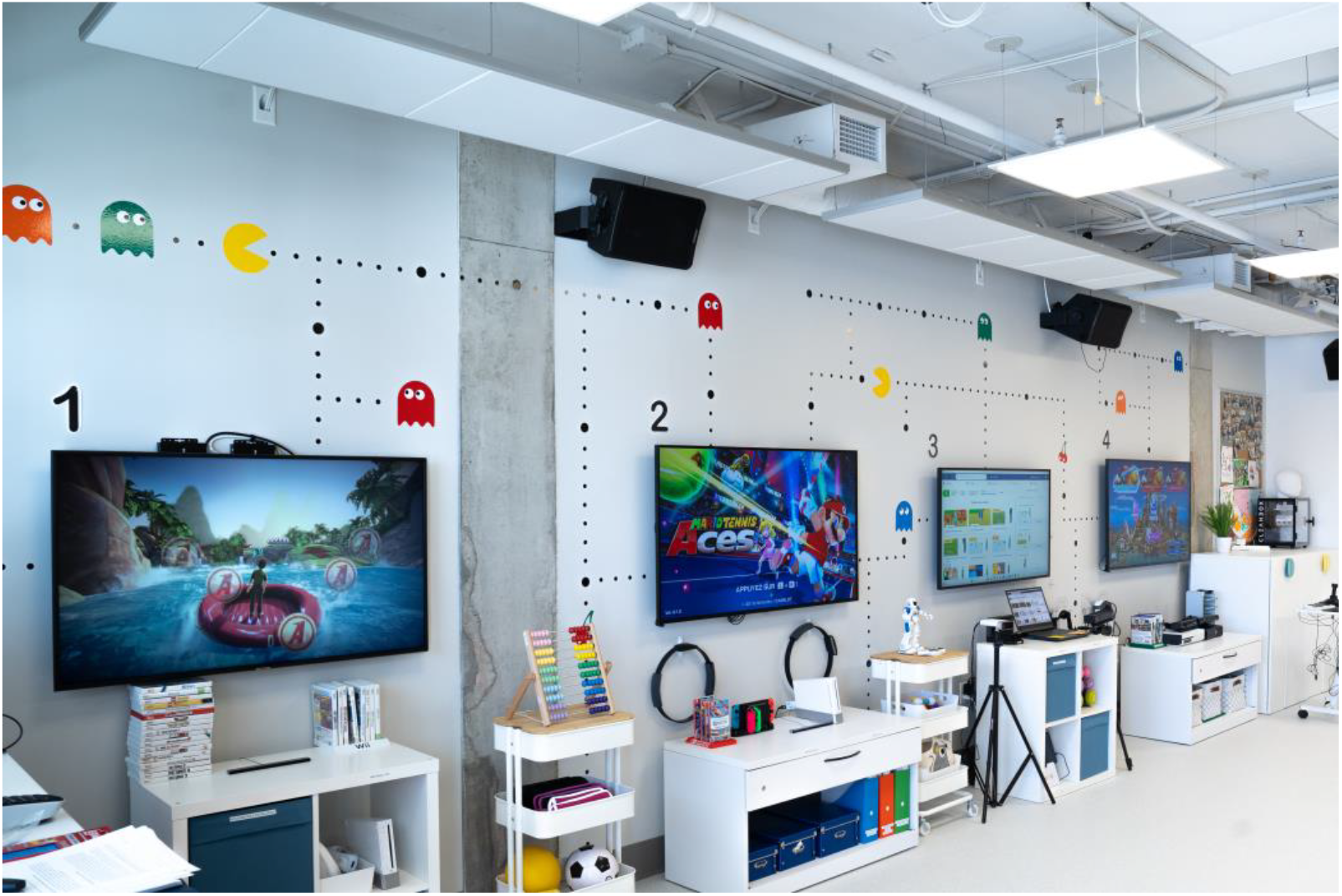
The Technotheque (© Studio 55 Productions)

## Objectives

The overall objective of this study is to assess the feasibility of the *Technotheque*, a knowledge mobilization initiative designed to provide access, training and support for VR/AVG use in one pediatric rehabilitation center. Feasibility will be evaluated through 4 sub-objectives based on key criteria suggested by Bowen and al. (2009) (47); each sub-objective will be evaluated using both quantitative and qualitative measures.

1. Demand is defined as the extent to which a new program is likely to be used. Demand will be quantified by the number of Technotheque visits and described by nature of VR/AVG use (e.g., systems, games, clinical objectives, and demographic characteristics of clinicians and clients).
2. Acceptability is defined as the extent to which a new program is judged as suitable, satisfying or attractive to recipients. Acceptability will be quantified by total score on a post-study satisfaction questionnaire and described in relation to Technotheque use during the study.
3. Adaptation is defined as how a program performs when changes are made. Adaptation will be described by the variety (or lack thereof) of Technotheque use in terms of professions, clients, and functional objectives.
4. Implementation is defined as the extent to which a new program can be successfully delivered to the intended participants in some defined, but not fully controlled context. Implementation will be quantified by pre-post study change in VR/AVG attitudes, facilitators and barriers, and described by the nature of Technotheque support personnel involvement in intervention sessions. Implementation feasibility will also be described via participant feedback on logistical features of Technotheque use such as the reservation system, the required paperwork, and the recruitment events.

## Methods

### Study design

This four-month study uses a convergent mixed-methods approach in which quantitative and qualitative data are collected simultaneously and then compared and/or combined for a comprehensive understanding (48).

### Setting

The *Technotheque* is located at the CRME’s Technopôle in Pediatric Rehabilitation in Montreal, QC. The CRME provides specialized rehabilitation services to children and youth aged 0-18 years with motor or speech disabilities. The CRME operates through six core programs: amputation and other musculoskeletal lesions, communication disorders, cerebral motor deficits, neuromuscular diseases, neuro-traumatology and motor development disorders. The inpatient unit provides intensive rehabilitation to children and youth recovering from traumatic injuries or planned surgeries. CRME services are provided by a team of approximately 130 rehabilitation professionals including physiotherapists, occupational therapists, speech therapists, special educators, and neuropsychologists.

## Participants

### Inclusion and exclusion criteria

All CRME clinicians who are interested in using the *Technotheque* will be invited to participate in the study. There are no exclusion criteria. There are approximately 130 clinicians at CRME; however, many work with infant and preschool-aged children and therefore would be less interested in using the Technotheque.

### Recruitment

We will use convenience and snowball (word of mouth) recruitment strategies. The study will begin with a one-week ‘open house’ event in which dedicated drop-in informational and hands-on trial sessions will be provided for VR/AVG use targeting different clinical goals (for example, balance, fine motor skills, or social participation). Given that clinicians can begin their participation at any point during the 4-month study, there will be weekly drop-in lunch time informational sessions offering hands-on trial opportunities throughout the study. In addition, a monthly e-newsletter will be sent will be sent out to all CRME clinicians, reminding them about the Technotheque and the study procedures.

## Material

**Appendix A** lists the VR/AVG systems and therapy equipment in the Technotheque.

### Study procedures

**Figure 2** outlines the 3 phases of the study procedures. Participants can begin and end their involvement at any point during the study. One of 3 trained Technotheque knowledge brokers (rehabilitation students) will be available during regular working hours for the 4-month study.

**Figure 2.**
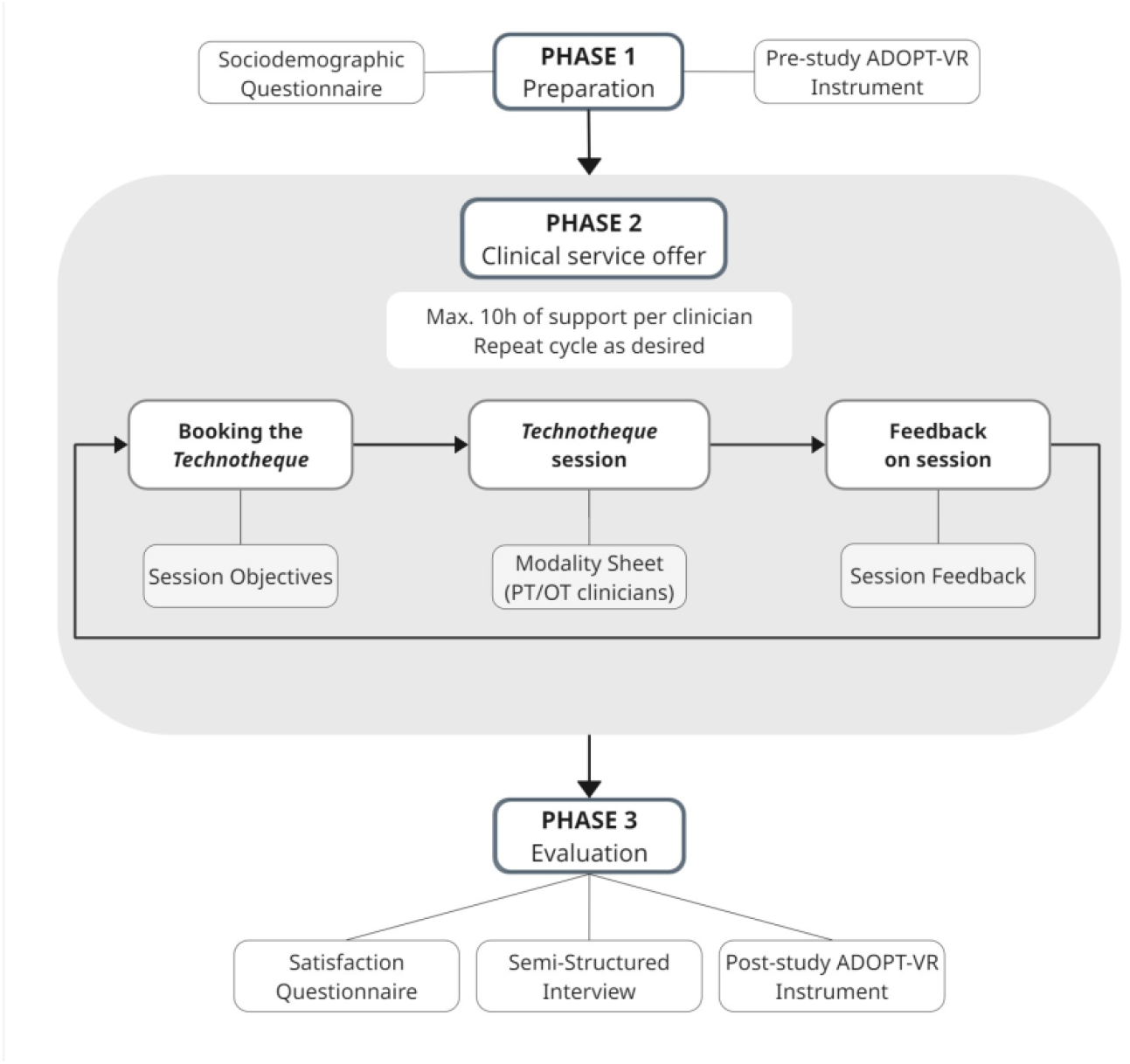
Study procedures

#### Phase 1: Preparation

Following informed consent, participants will complete a sociodemographic questionnaire and the Assessing Determinants of Prospective Take Up of Virtual Reality (ADOPT-VR2) instrument before their first Technotheque use. Children and youth and their parents will not be required to sign an assent/consent form for Technotheque use in the context of this study, as clinicians are choosing to use the Technotheque within their regular service offer. However, there will be an option for youth and for parents to participate in a short interview regarding their Technotheque experience. Knowledge brokers will complete informed assent/consent procedures with youth participants and their caregivers.

#### Phase 2: Clinical and educational support services in the Technotheque

Clinician participants or the study coordinator will reserve the Technotheque for each session using the CRME reservation system. The coordinator, who monitors the reservation calendar, will email the pre-session objectives form for completion by the clinician. Participants can reserve 1 of 2 types of individualized assistance: 1) A therapy session occurring in a clinical context during which the knowledge broker offers game selection and implementation support as well as technical assistance throughout the session. Participants self-select the in-patient or out-patient clients that they choose to bring to the Technotheque, as well as the frequency of Technotheque use; or 2) An educational session outside of a clinical context in which the knowledge broker provides educational or technical training, for a maximum of 10 hours of paid support. Following each session, the study coordinator will send out the post-session feedback form. Physiotherapists and occupational therapists will also complete a custom modality sheet detailing VR/AVG system and game use; this form is designed to be included in their charting process.

In consultation with the knowledge broker, participants can decide at any point when they are ready to move to independent clinical use of the Technotheque. This decision can be specific to use of a certain system, and participants can again request assistance with use of a different VR/AVG system. Even while independent, clinicians will be asked to complete the pre- and post-session forms.

#### Phase 3: Evaluation

At the end of the study, or when an individual participant decides to end their participation at any point during the study, they will be asked to complete the post-study ADOPT-VR2, a CFIR-based study-specific satisfaction questionnaire, and a semi-structured interview.

## Data collection measures

### Sociodemographic questionnaire

Describes clinicians’ professional profiles, years of clinical experience, clinical populations with whom they work, and prior personal and professional VR/AVG experience.

### Assessing Determinants of Prospective Take-up of Virtual Reality (ADOPT-VR) Instrument

The ADOPT-VR2 is designed to assess clinicians’ attitudes and perceptions regarding VR adoption in rehabilitation (40). The instrument has established face and content validity, and high internal consistency (Cronbach’s alpha=0.876) as well as demonstrated responsiveness (40,41). The instrument was modified to suit our context, by removing options related to adult patients. The survey consists of 54 items derived from the Theory of Planned Behavior, assessing theoretical predictors of behavioral intention to use VR. The constructs are grouped into three composites: (1) Attitudes: Perceived Usefulness, Perceived Ease of Use, and Compatibility; (2) Social Norms: Peer Influence, Superior Influence, Client Influence; and (3) Perceived Behavioral Control: Self Efficacy, Facilitating Conditions and Barriers. In this study, we will focus on CO, CI, and FCB constructs, with item-level analyses for specific items within the remaining constructs.

### Planning and Documentation of Technotheque use

Three study-specific forms will capture information related to Technotheque use. 1) Session Objectives questionnaire: Prior to each therapy session, participants will complete a short online questionnaire consisting of multiple-choice responses and open-ended questions indicating therapy objectives, reasons for using the Technotheque, and providing relevant client information (for example, functional capacities and interests/hobbies). 2) Session Feedback questionnaire: This form combines multiple choice questions and agreement questions on a 7-point Likert scale (with anchors on strongly disagree and strongly agree) documenting clinician satisfaction with each session. The form also provides space to list technical issues or equipment needs. 3) Modality Sheet: Specific to physiotherapists and occupational therapists, this form was created to facilitate documentation of game use to target specific objectives during a therapy session, with the intention of supporting charting activities.

### Post-study satisfaction questionnaire

We developed a CFIR-based questionnaire with 56 items distributed across 4 CFIR domains (Innovation, Inner Setting, Individuals, Implementation). Feasibility analyses we will focus on satisfaction-specific questions specific in the Innovation domain (e.g., clinical relevance of available technologies, perceived ease of use, relative advantage of in-person support) and the Inner Setting domain (e.g., compatibility of the service offer with existing administrative procedures, adequacy of the physical infrastructure); these questions can be found in **Appendix B**. Internal consistency will be assessed using Cronbach’s alpha. With sufficient consistency, a total satisfaction score will be used for analyses. Findings from the remaining questions unrelated to satisfaction will be considered alongside qualitative findings in the mixed methods analysis.

### Semi-structured interview

Will be conducted at the end of a clinician’s study participation by a research coordinator. The interview guide was developed by the study team to further explore and expand upon CFIR-based satisfaction questionnaire. **Appendix C** lists the questions designed to probe clinicians’ perspectives on facilitators and barriers to Technotheque use. Analyses of youth and parent interviews will be the focus of a subsequent publication and will not be included in feasibility analyses.

## Analyses

### Quantitative analyses

Quantitative analyses will be conducted using IBM SPSS Statistics 27, with Shapiro-Wilk tests employed to assess normality of data distributions. Clinician demographic data will be summarized using descriptive statistics.

#### Objective 1

Demand will be quantified by descriptive statistics summarizing participant demographics, nature of VR/AVG use (e.g., types of VR/AVG systems used, clinical objectives, types of clients), and the number of visits per participant, transformed to percentiles and categorized as low (below 25^th^ percentile), medium (between 25^th^ and 75^th^ percentile), or high (above 75^th^ percentile). Chi-square analyses will be conducted to investigate the relationships between users’ demand levels (low, medium, or high) and demographic variables such as pre-study professional or personal use of VR/AVGs and client profile (in-patient vs out-patients).

#### Objective 2

Acceptability will be quantified by descriptive statistics summarizing post-session feedback questionnaires as well as the post-study Satisfaction questionnaire total score. Mean satisfaction scores will be compared across user demand levels (low, medium, high) using a one-way ANOVA or a Kruskal-Wallis test.

#### Objective 3

Adaptation will be quantified by descriptive statistics that indicate the variety in Technotheque use across client characteristics (sex, age groups, clinical program, functional mobility status and therapeutic objectives).

#### Objective 4

Implementation will be assessed by parametric or non-parametric within-group pre-post differences in ADOPT-VR2 scores. To control for pre-study Technotheque exposure (i.e., participation in the pilot phase), analyses will exclude clinicians who participated in the pilot phase of Technotheque development. Implementation feasibility will also be quantified through descriptive statistics and visual analyses of nature of Technotheque use over the four-month period with respect to bookings with the knowledge broker vs independent Technotheque use.

### Qualitative analyses

Semi-structured interviews will be transcribed and analyzed in NVivo 15 using Bowen’s feasibility criteria as a predefined coding framework. Any responses to open-ended questions in the post-study ADOPT-VR and the Satisfaction questionnaire will be added to the qualitative data. The qualitative analysis method will follow a 4-step iterative ‘critical friends’ approach, a collaborative and reflective process where researchers critically engage with one another’s data interpretations.(49)

#### Step 1: Primary Deductive Analysis

Two investigators (AF, ED) will independently analyze a randomly selected 25% of the transcripts to compare coding approaches, discuss similarities and differences, and iteratively refine the coding framework.

#### Step 2: Critical Appraisal and Feedback

Two additional study team members will familiarize themselves with Bowen’s feasibility criteria and review the same initial 25% of transcripts. AF will then present the preliminary coding and interpretations during a critical appraisal session, where team members will provide constructive feedback. This session will serve as a theoretical sounding board, promoting reflection and encouraging alternative interpretations of the data.

#### Step 3: Refinement and Iteration

Following the feedback session, AF will revise the coding framework and interpretations based on the group’s discussions and consensus. This process will be repeated with the next set of five transcripts, ensuring ongoing refinement and enhanced rigor throughout the analysis. The cycle will continue until all qualitative data has been analyzed.

#### Step 4: Member Checking

To enhance the trustworthiness of the findings, the study team will invite participants to an optional member-checking discussion where they will have the opportunity to provide feedback on the interpretations (50).

### Mixed-methods analyses

The integration of quantitative and qualitative data will follow the procedures outlined in the convergent design framework (47): 1) Quantitative and qualitative data will first be analyzed separately; 2) Common concepts will be derived each of the datasets; 3) A joint display (e.g. a table or graph) will be developed to facilitate comparisons; 4) Results will be compared to determine in what ways they confirm, disconfirm or expand each other; and 5) Interpretations will be finalized by the study team and presented in the context of the 5 CFIR domains.

## Results

This study was approved by the CHU Sainte-Justine research ethics committee.

## Discussion

This study will use a convergent mixed-methods approach to explore the feasibility of a multifaceted knowledge mobilization initiative targeting access and educational barriers to the use of VR/AVG systems at a pediatric rehabilitation centre. Although the study is limited to one clinical site, our findings will contribute to the evidence base on the factors that support or limit clinical use of VR/AVGs in pediatric rehabilitation and can identify facilitators and barriers to inform the development of similar initiatives at other sites. Study results will inform the direction of subsequent research directions exploring the potential of the Technotheque as a knowledge translation strategy and the clinical effectiveness of VR/AVG use as a rehabilitation intervention.

## Data Availability

There is no data related to this manuscript.

## Appendix A

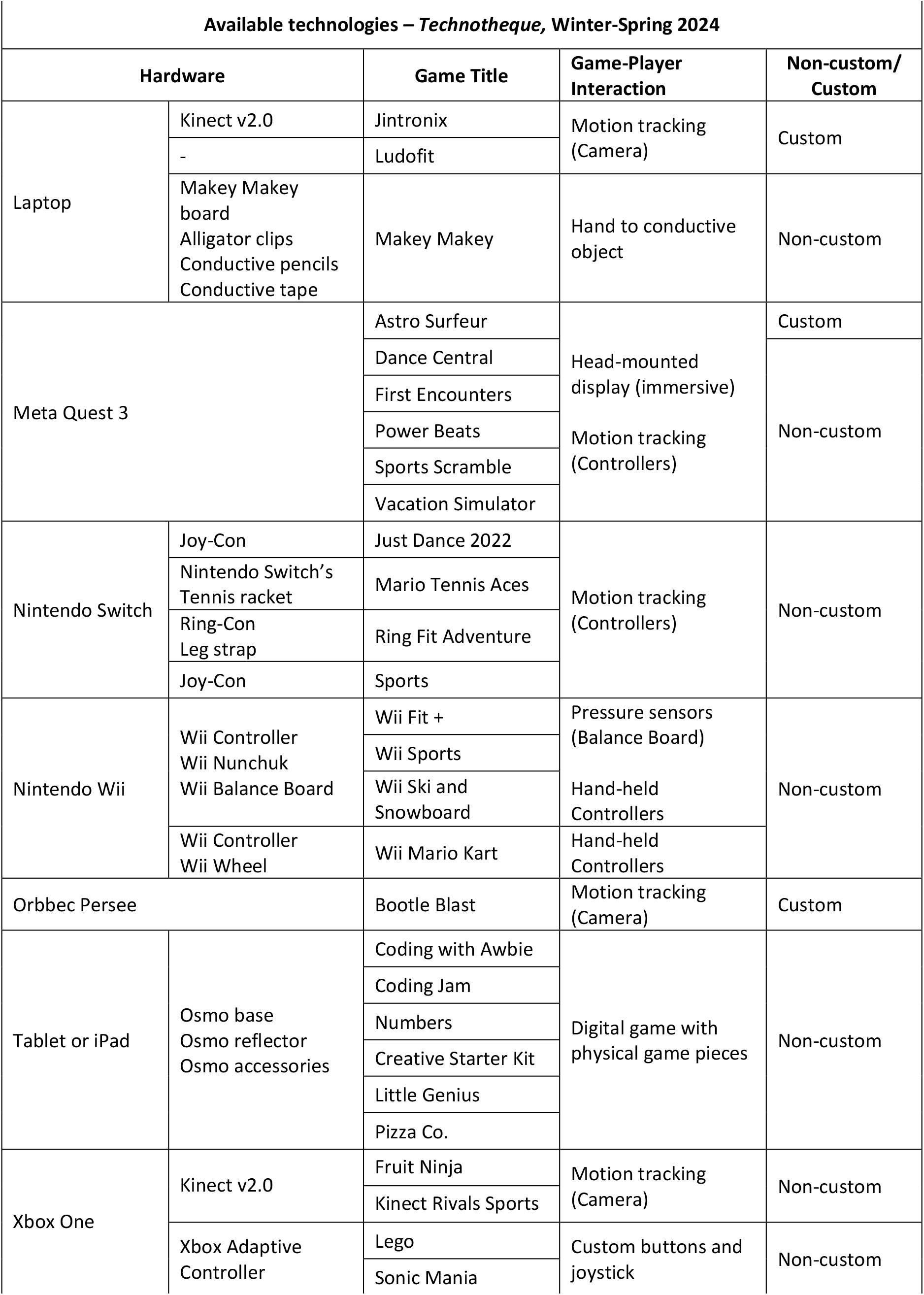
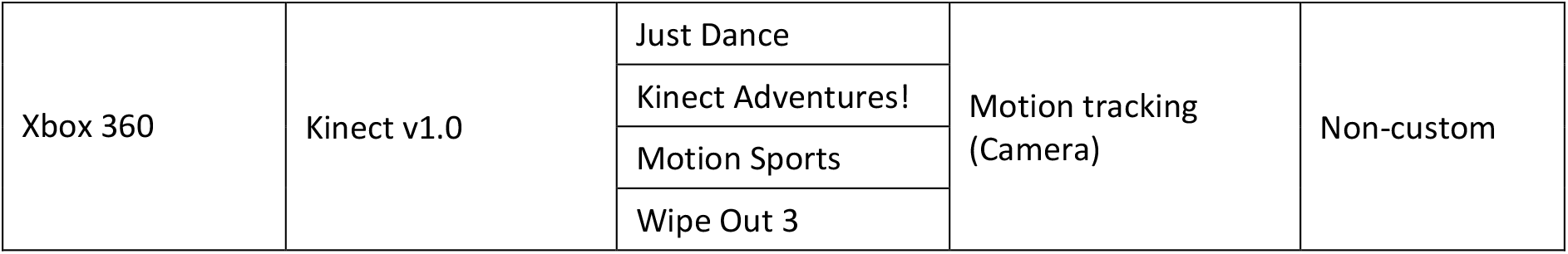

## Appendix B

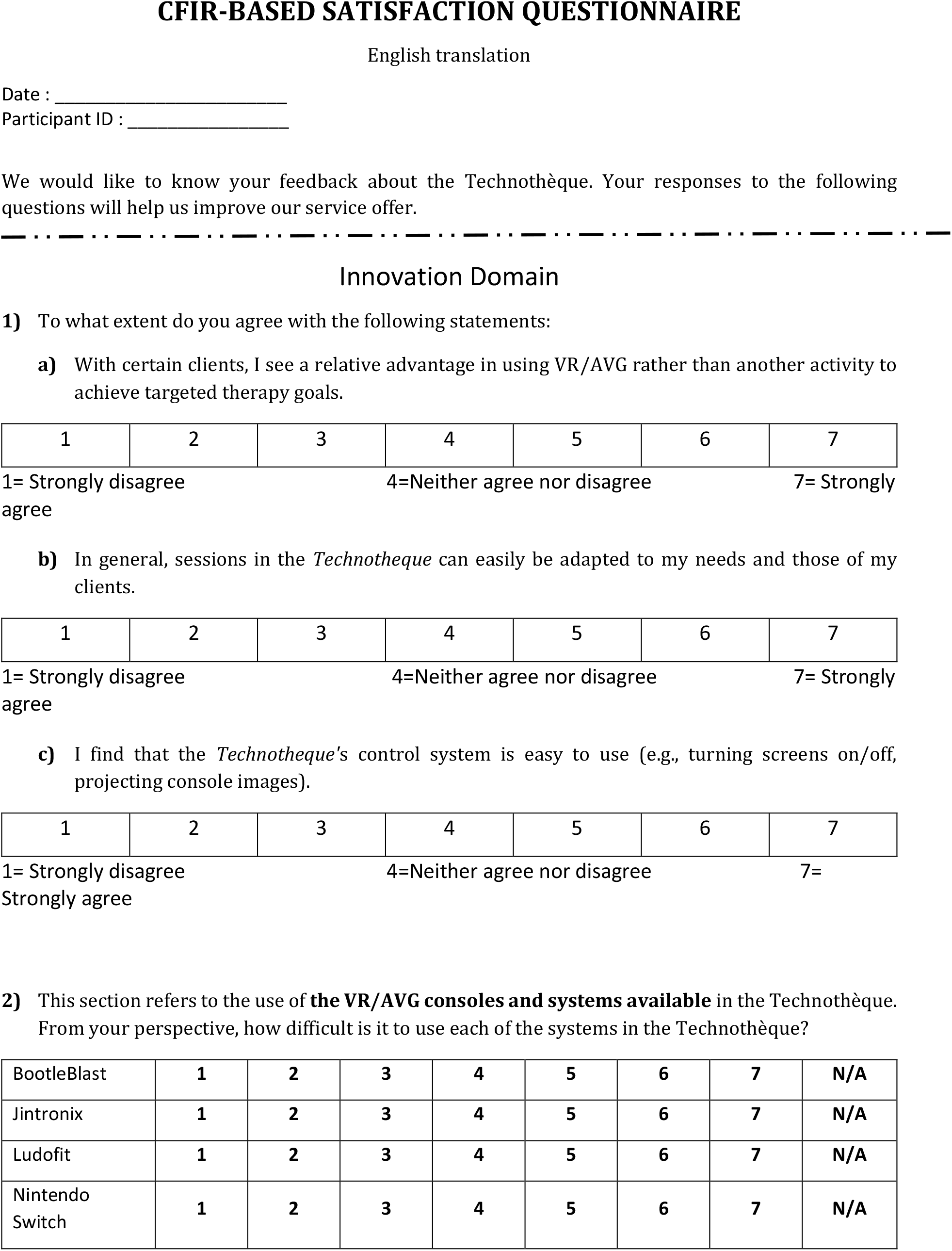
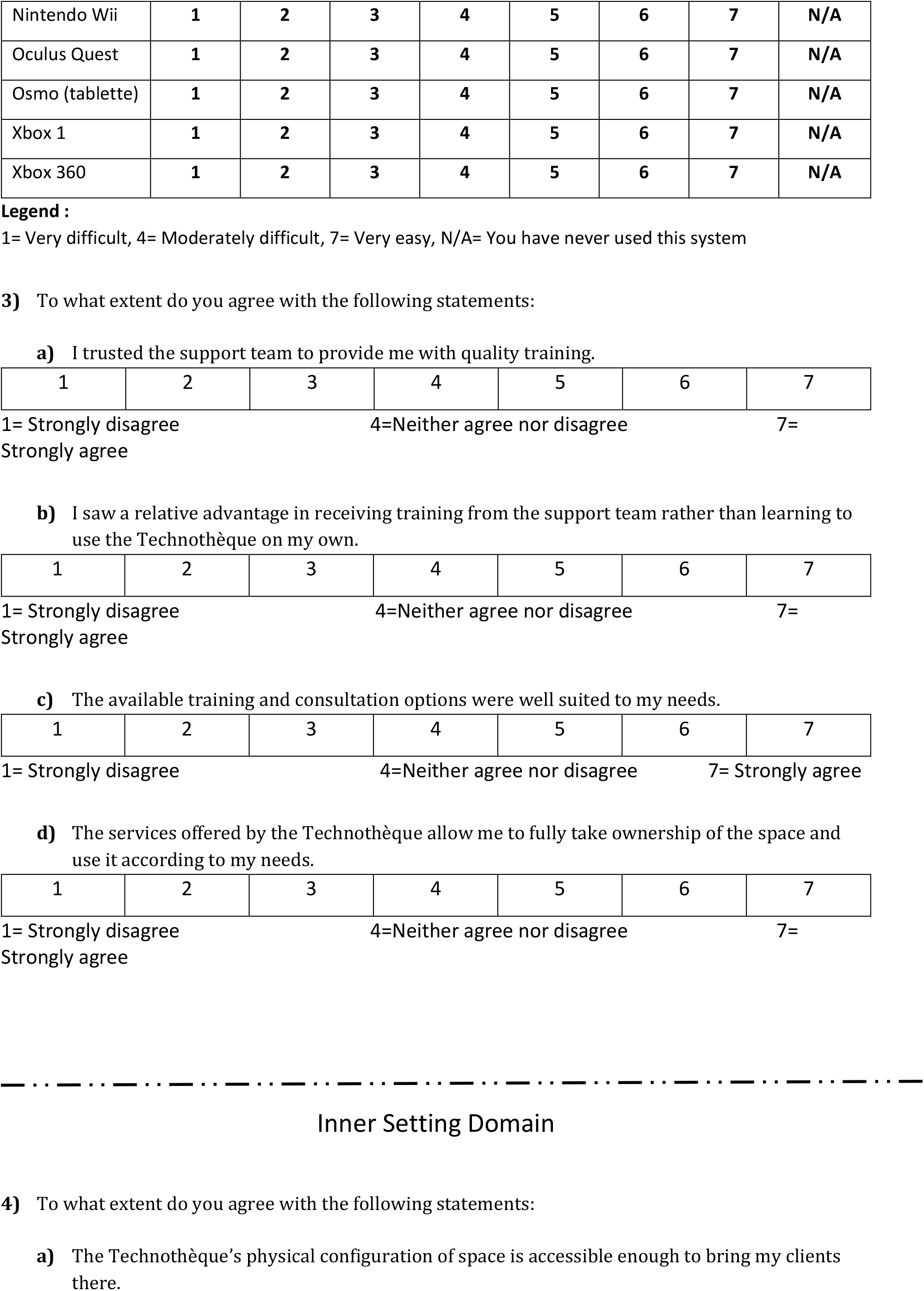
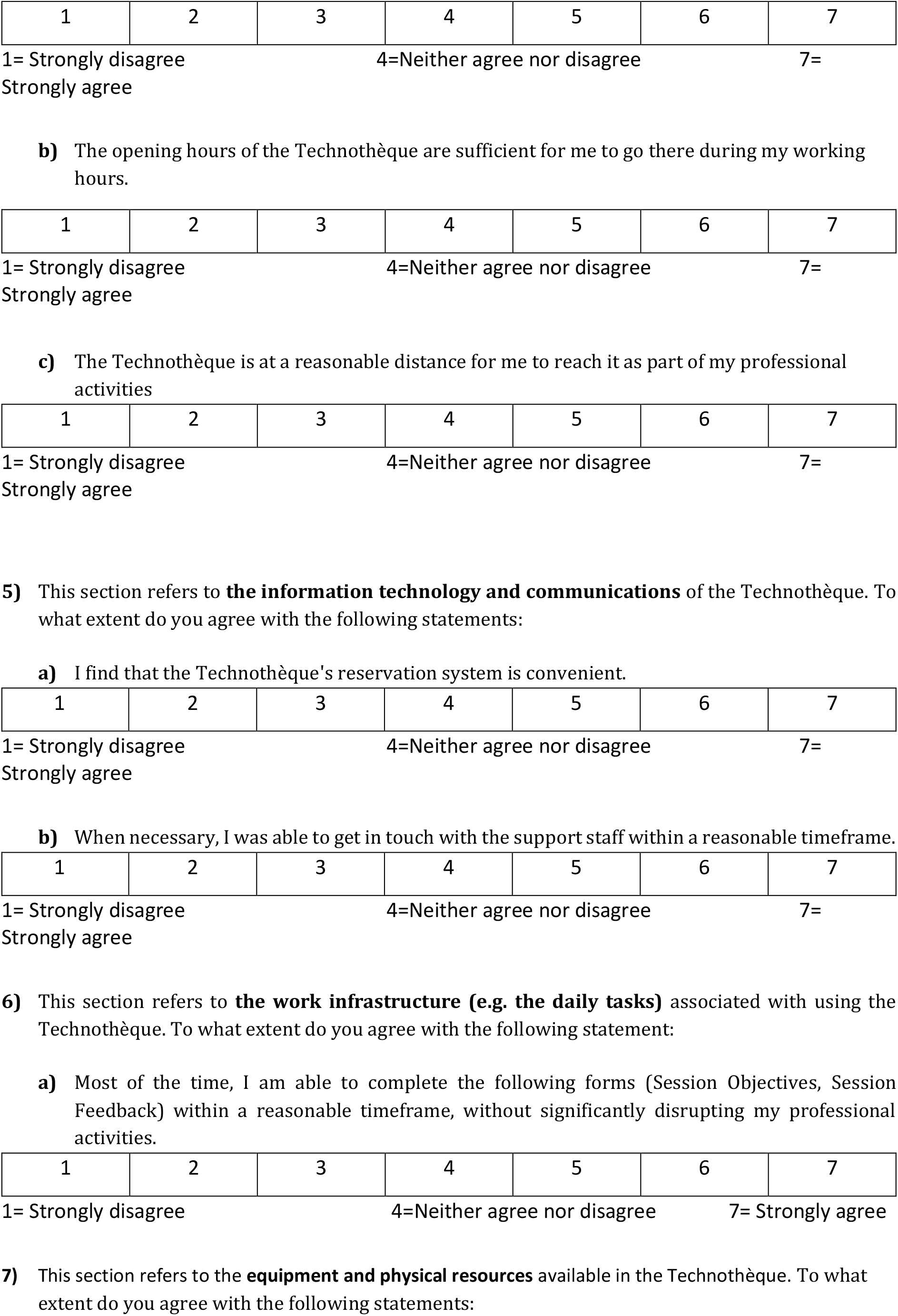
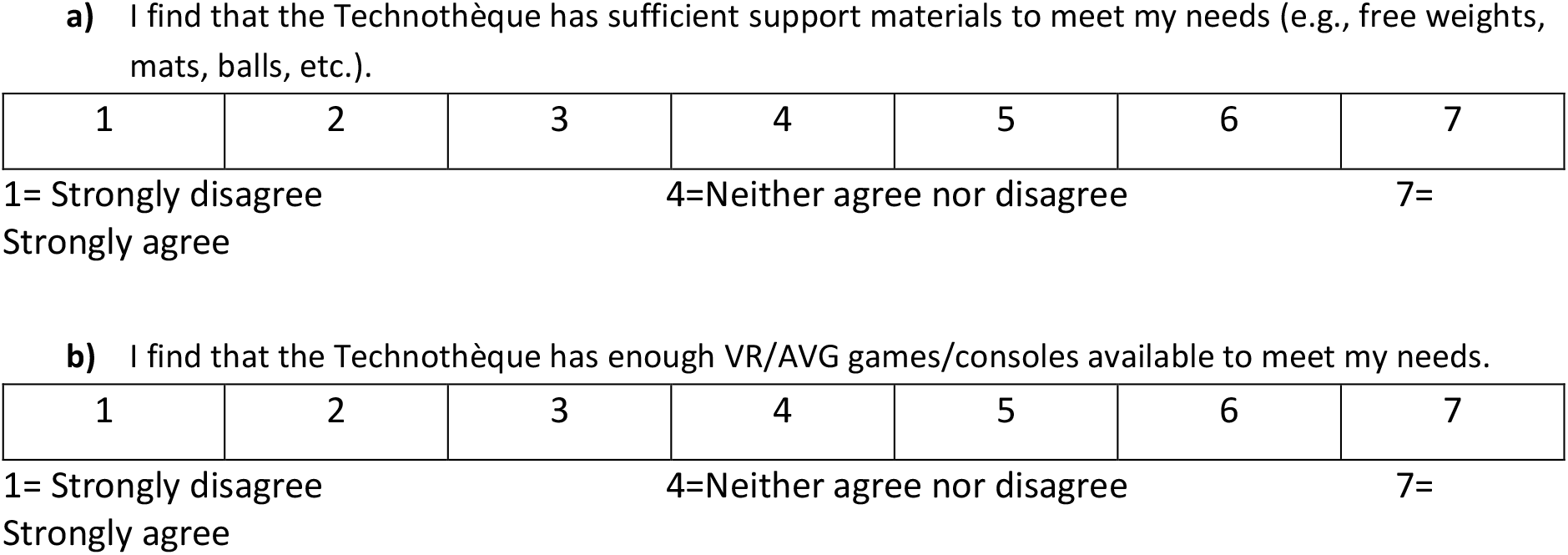

|  |  |  |  |  |  |  |
|---|---|---|---|---|---|---|
| 1 | 2 | 3 | 4 | 5 | 6 | 7 |
| 1 | 2 | 3 | 4 | 5 | 6 | 7 |
| 1 | 2 | 3 | 4 | 5 | 6 | 7 |

|  |  |  |  |  |  |  |  |  |
| --- | --- | --- | --- | --- | --- | --- | --- | --- |
| BootleBlast | 1 | 2 | 3 | 4 | 5 | 6 | 7 | N/A |
| Jintronix | 1 | 2 | 3 | 4 | 5 | 6 | 7 | N/A |
| Ludofit | 1 | 2 | 3 | 4 | 5 | 6 | 7 | N/A |
| Nintendo Switch | 1 | 2 | 3 | 4 | 5 | 6 | 7 | N/A |
| Nintendo Wii | 1 | 2 | 3 | 4 | 5 | 6 | 7 | N/A |
| Oculus Quest | 1 | 2 | 3 | 4 | 5 | 6 | 7 | N/A |
| Osmo (tablette) | 1 | 2 | 3 | 4 | 5 | 6 | 7 | N/A |
| Xbox 1 | 1 | 2 | 3 | 4 | 5 | 6 | 7 | N/A |
| Xbox 360 | 1 | 2 | 3 | 4 | 5 | 6 | 7 | N/A |
| **a) I trusted the support team to provide me with quality training.** |  |  |  |  |  |  |
| 1 | 2 | 3 | 4 | 5 | 6 | 7 |
| 1 | 2 | 3 | 4 | 5 | 6 | 7 |
| **c) The available training and consultation options were well suited to my needs.** |  |  |  |  |  |  |
| 1 | 2 | 3 | 4 | 5 | 6 | 7 |
| 1 | 2 | 3 | 4 | 5 | 6 | 7 |
| **a) The Technothèque's physical configuration of space is accessible enough to bring my clients there.** |  |  |  |  |  |  |
| 1 | 2 | 3 | 4 | 5 | 6 | 7 |
| 1 | 2 | 3 | 4 | 5 | 6 | 7 |
| 1 | 2 | 3 | 4 | 5 | 6 | 7 |
| 1 | 2 | 3 | 4 | 5 | 6 | 7 |
| 1 | 2 | 3 | 4 | 5 | 6 | 7 |
| 1 | 2 | 3 | 4 | 5 | 6 | 7 |
| 1 | 2 | 3 | 4 | 5 | 6 | 7 |
| 1 | 2 | 3 | 4 | 5 | 6 | 7 |

## Appendix C

### SEMI-STRUCTURED INTERVIEW GUIDE

1. Could you tell me a little bit about your use of the *Technotheque* during this research project?
2. What are your impressions of this experience?
  i. What did you like about your experience? Could you provide a specific example?
  ii. What did you not like about your experience? Could you provide a specific example?
3. From your perspective, what might be the benefits, if any, of the *Technotheque* for your clinical practice?
4. In your opinion, are there specific profiles of children or youth for whom the *Technotheque* would be particularly relevant? Examples might include children of certain ages, specific physical/cognitive disabilities, personality traits, or interests.
  i. Conversely, are there profiles children or youth for whom the *Technotheque* would not be relevant?
5. (**If applicable**) Which game, if any, was particularly useful during your therapy sessions with a patient?
  i. Why?
6. (**If applicable**) Which game, if any, did you particularly enjoy for promoting social participation and/or enjoyment?
  i. Why?
7. (**If applicable**) What did you think of the *Technotheque’s* educational service offer?
  i. Do you have any suggestions for improvement?
8. (**If applicable**) What did you think of the Technotheque’s therapy support service offer?
  i. Do you have any suggestions for improvement?
9. Do you have suggestions for equipment we should acquire to support your activities in the *Technotheque*?
10. Do you have suggestions for systems or games we should acquire to better meet your needs?
  i. If ‘anything were possible,’ what would you like to be able to do with video games in the *Technotheque*?
11. In your opinion, what are the next priority activities that the *Technotheque* should organize? Examples might include group training sessions, development of new games, improving current services, etc.
12. Is there any other information that you would like to share with us?

